# Cost-effectiveness of antenatal screening for pre-eclampsia using PlGF and sFlt-1/PlGF ratio in Tanzania: A pre-trial health economic modelling study

**DOI:** 10.64898/2026.05.04.26352426

**Authors:** Giorgia Lovecchio, Andrea Solnes Miltenburg, Richard Kiritta, Albert Kihunrwa, Anne Cathrine Staff, Lumbwe Chola

**Affiliations:** Department of Health Management and Health Economics, University of Oslo, Oslo, Norway; Department of Community Medicine and Global Health, Institute of Health and Society, Faculty of Medicine, University of Oslo, Oslo, Norway; Department of Obstetrics and Gynaecology, Akershus University Hospital, Lørenskog, Norway; Department of Obstetrics and Gynecology, Bugando Medical Centre, Catholic University of Health and Allied Sciences, Mwanza, Tanzania; Institute of Clinical Medicine, Faculty of Medicine, University of Oslo, Oslo, Norway; Division of Obstetrics and Gynaecology, Oslo University Hospital, Oslo, Norway

**Keywords:** pre-eclampsia, biomarker screening, PlGF, sFlt-1/PlGF, cost-effectiveness analysis, Tanzania

## Abstract

Pre-eclampsia (PE) is a major contributor to maternal and neonatal morbidity and mortality. Research in high-income countries has shown that biomarker-based PE screening can improve timely detection and management of women at high risk of PE. We conducted a pre-trail cost-effectiveness analysis of introducing biomarker screening in Tanzania to identify key parameters informing a full, trial-based economic evaluation. We developed a decision tree comparing current practice with two biomarker-based screening strategies: Strategy 1 introducing placental growth factor (PlGF), and Strategy 2 adding soluble fms-like tyrosine kinase-1 (sFlt-1)/PlGF ratio. Under both strategies we assumed early aspirin prophylaxis and/or close monitoring for high-risk women. For each of the three options, we modelled PE diagnosis, as well as maternal and neonatal outcomes over a one-year time horizon, assuming a healthcare sector perspective. We quantified health outcomes in terms of disability-adjusted life years (DALYs) and costs in 2023 US$.

When compared to current practice, the incremental cost per DALY averted was $410.45 for Strategy 1 and $1,011.78 for Strategy 2. Limiting the novel strategies to nulliparous women, decreased incremental cost-effectiveness ratios to $184.15 (Strategy 1) and $413.33 (Strategy 2). Key parameters impacting cost-effectiveness were PE prevalence, biomarker screening accuracy, adherence to and effectiveness of preventive treatment and monitoring, and related costs.

Based on our findings, biomarker screening has the potential to be cost-effective in Tanzania, particularly if introduced early in pregnancy and targeted at nulliparous women. Further research in low-resource settings is needed to overcome the current data and evidence gaps.

## Introduction

Pre-eclampsia (PE), a hypertensive disorder of pregnancy, affects approximately 3-5% of pregnancies worldwide. Every year, an estimated 76,000 maternal deaths, 500,000 perinatal deaths and 200,000 stillbirths are associated with PE [1,2]. Most of these birth complications and deaths occur in low- and middle-income countries (LMICs) [2]. In sub-Saharan Africa, hypertensive disorders account for 16% of maternal deaths [3]. In Tanzania, the prevalence of PE and eclampsia is 4.2%, contributing to approximately 22% of maternal deaths, 23% of maternal near misses (MNMs), and 41% of stillbirths [4,5].

If left untreated, PE can lead to severe maternal complications, such as eclampsia, HELLP syndrome, stroke, maternal death, and increased long-term cardiovascular risk. For the offspring, untreated PE may lead to foetal death, preterm birth, growth restriction, and neurodevelopmental disorders [1]. Early screening and timely management of high-risk women are crucial for improving maternal and neonatal outcomes. Known maternal risk factors for PE include primiparity, pre-existing hypertension, chronic kidney disease, diabetes mellitus, previous PE, obesity and family history of PE [1]. In most LMICs, including Tanzania, antenatal screening for PE remains predominantly opportunistic and risk-factor-based. However, this approach has limited predictive value, failing to identify a substantial portion of high-risk women [1]. This limitation reflects the complex aetiology of PE, incomplete understanding of its pathophysiology, heterogeneous clinical manifestations, and differences across populations [6].

Clinical guidelines now recommend incorporating placenta-associated biomarkers into early pregnancy screening to improve prediction accuracy [1,7]. When combined with maternal demographic characteristics and blood pressure measurements, biomarkers can aid in identifying pregnant women who may benefit from intensified surveillance or early therapeutic interventions [8]. Key biomarkers include placental growth factor (PlGF), a proangiogenic protein often reduced prior to clinical PE diagnosis, and antiangiogenic soluble fms-like tyrosine kinase-1 (sFlt-1), another placenta-associated protein, often elevated before and during PE. In the second half of pregnancy, their combined measurement is expressed as the sFlt-1/PlGF ratio, with higher values indicating greater disease risk [8]. In first trimester screening, PlGF remains the most efficient biomarker for prediction algorithms [9]. Economic evaluation studies have shown that incorporating such biomarkers into clinical practice can be highly cost-effective [10–12].

Most studies of biomarker screening for PE come from high-income countries (HICs), leading to the development of complex multi-parameter laboratory assays utilising proteomic and metabolomic analytical techniques. These techniques may not be practical in low-resource settings due to limited laboratory infrastructure and prohibitive costs [13]. Further research is needed to evaluate the suitability, performance, and economic impact of biomarker-based PE screening in LMICs [13].

This study presents a pre-trial economic evaluation of the PREventing Severe Hypertensive Adverse events in pregnancy and childbirth (PRESHA) study, currently underway in Tanzania, to demonstrate the feasibility and efficacy of introducing PlGF and/or sFlt-1/PlGF ratio for PE screening in a low-resource setting [14]. To our knowledge, this is the first planned cost-effectiveness analysis of biomarker-based PE screening in sub-Saharan Africa. The study will help establish key assumptions and identify data to be collected in the planned trial, to enable a comprehensive and robust cost-effectiveness analysis of PE screening interventions.

## Methods

### Study Design

We developed a decision tree model, using Microsoft Excel (Version 16.94), to assess the cost-effectiveness of PE screening strategies introducing PlGF and sFlt-1/PlGF ratio measurement compared to current practice. The population was a hypothetical cohort of 3,000 Tanzanian women aged 16 or older, with a confirmed gestational age of less than 20 weeks. The base-case analysis adopts a healthcare sector perspective which accounts for all medical costs within the formal healthcare system, irrespective of the payer. A one-year time horizon is applied, covering pregnancy and three months postpartum to capture potential adverse outcomes. Given the limited time perspective, both costs and effects are not discounted. The 2022 CHEERS checklist was used to align with current reporting standards for health economic evaluations (S1 Table).

In this analysis PE is defined as new-onset hypertension at or after 20 weeks of gestation, coupled with new-onset proteinuria and/or other signs of maternal end-organ or uteroplacental dysfunction [2]. PE is subclassified as preterm (delivery <37^+0^ weeks) or term (delivery ≥37^+0^ weeks) PE [2]. Given the variation in prevalence and screening performance with respect to preterm and term PE, the analysis is conducted for these two sub-populations.

### Study Context

As part of the PRESHA project, the study is set in Mwanza, Tanzania, where antenatal care (ANC) services are government-funded, but considerable out-of-pocket payments are incurred for medication, delivery, and management of complications. In low-resource settings like Tanzania, ANC is typically initiated later than in HICs, and the quality of care is limited by restricted access to clinical laboratories and trained personnel [13]. Although 90% of pregnant women in Tanzania attend at least one ANC visit, only 65% complete four or more visits, and about 34% start ANC before 16 weeks of gestation [15]. National ANC guidelines, require a comprehensive physical examination and documentation of the medical and obstetric history of every pregnant woman, ensuring the identification of maternal risk factors for PE [16]. However, in the absence of specific routine screening for PE and subsequent risk stratification, women with identified risk factors are managed according to routine ANC protocols. PE is often diagnosed only upon the emergence of clinical signs later in pregnancy.

### Comparators: Biomarker-based tests

Two biomarker-based PE screening strategies are evaluated in this study. These strategies stratify women into high and low/no risk categories for PE, based on different screening methods and timing. Strategy 1 combines the identification of maternal risk factors, maternal mean arterial pressure (MAP) and levels of PlGF before 16 weeks of gestation. This approach is expected to improve the accuracy of PE prediction and prevent preterm PE through timely management of high-risk women. In line with international guidelines, management consists of early initiation of low-dose aspirin and close monitoring of high-risk women [17]. Following Zakiyah et al., (2022), close monitoring is quantified as four additional ANC visits and two ultrasounds.

Strategy 2 builds upon current practice by introducing MAP and sFlt-1/PlGF ratio measurements from 24 weeks of gestation, reflecting the physiological rise in sFlt-1 levels starting around 21–24 weeks [8]. This strategy aims to identify high-risk women later in pregnancy to prevent adverse outcomes through close monitoring. Low-dose aspirin is not recommended, as it is ineffective when started at this stage of gestation [18].

### Modelling

The decision tree model, illustrated in Fig 1, aligns with previous cost-effectiveness analyses on PE screening strategies [11,19–21]. At the decision node, three options are compared: current practice, Strategy 1, and Strategy 2. The primary outcome of each screening option is PE diagnosis, which is categorised as PE <37 (preterm PE), PE ≥ 37 (term PE), and No PE. For each primary outcome, the model tracks maternal and neonatal outcomes. Maternal outcomes include maternal death, MNM, and no complications. MNMs are defined as women who survive potentially life-threatening conditions during pregnancy, childbirth or postpartum [22]. In this analysis, MNM is used as a proxy for severe maternal complications arising from PE. Neonatal outcomes include live birth, preterm or at term, and stillbirth. Adverse outcomes such as low birth weight, NICU admission, low APGAR score, and neonatal death, are correlated to preterm birth. The interdependence of these outcomes makes individual modelling within a decision tree challenging. We therefore assume that preterm birth alone can encompass multiple sources of perinatal morbidity and mortality.

**Fig 1.**
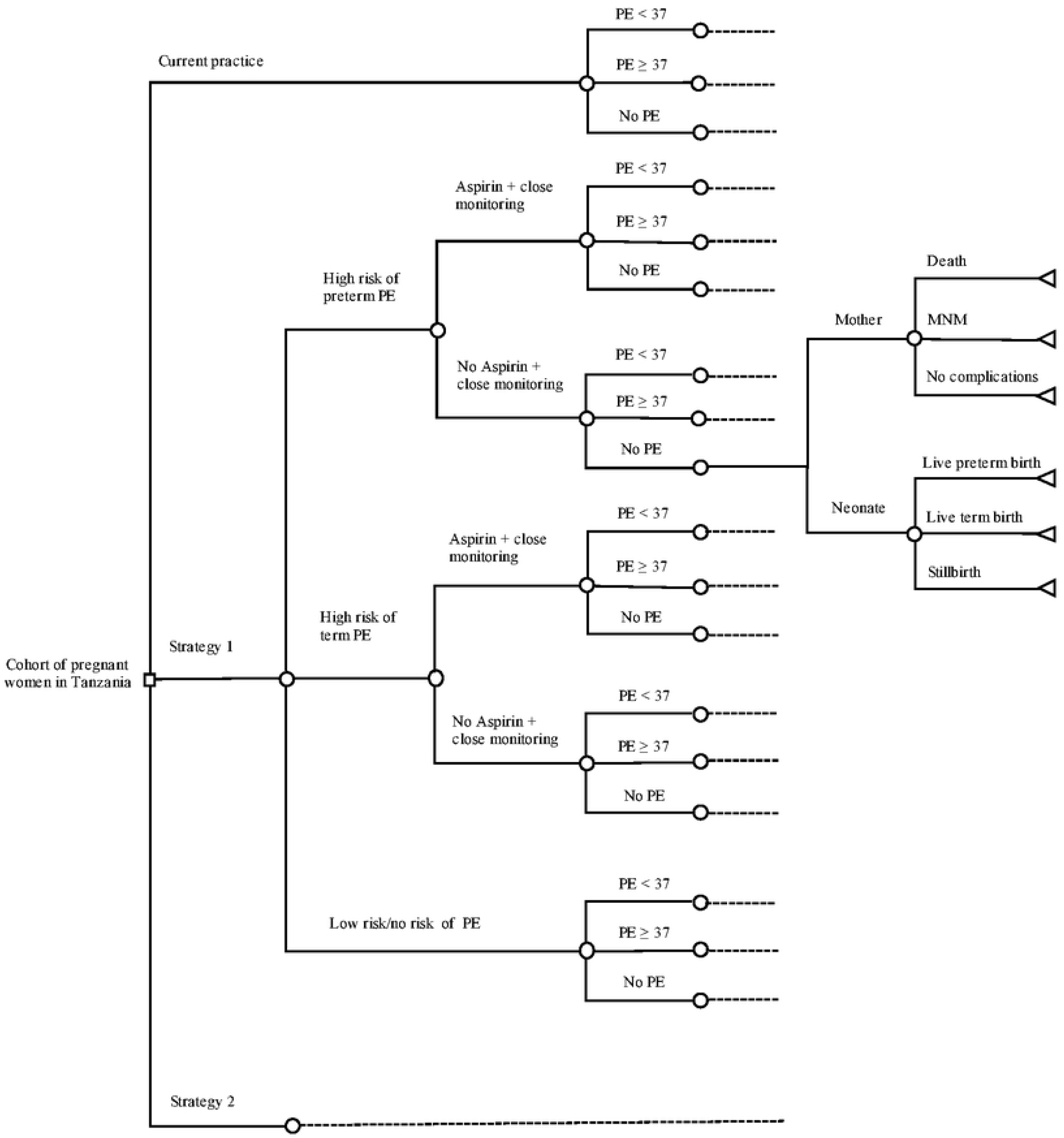
Decision tree model comparing current practice with two biomarker-based PE screening strategies. Current practice consists in the identification of maternal risk factors. No risk stratification nor preventive measures for high-risk women follow. Strategy 1 adds measurements of mean arterial pressure (MAP) and Placental growth factor (PlGF) levels to current practice, as well as risk stratification, aspirin and close monitoring for high-risk women. No aspirin + close monitoring refers to non-adherence to preventive recommendations. Each PE diagnosis decision node is followed by related maternal and neonatal outcomes. Strategy 2, adding MAP and soluble fms-like tyrosine kinase-1 (sFlt-1)/PlGF ratio measurements, unfolds in the same way as Strategy 1, except for managing high-risk women through close monitoring only, no aspirin. MNM = Maternal near miss. PE = Pre-eclampsia. PE<37 = Preterm PE. PE≥37 = Term PE.

### Data Inputs

Local data and estimates from Tanzania or comparable low-resource settings were prioritised, with HIC data used only when necessary and verified for applicability. The PRESHA team at Bugando Medical Centre helped refine model assumptions and validate data inputs. Model parameters, values, and sources are summarised in Table 1.

**Table 1.**
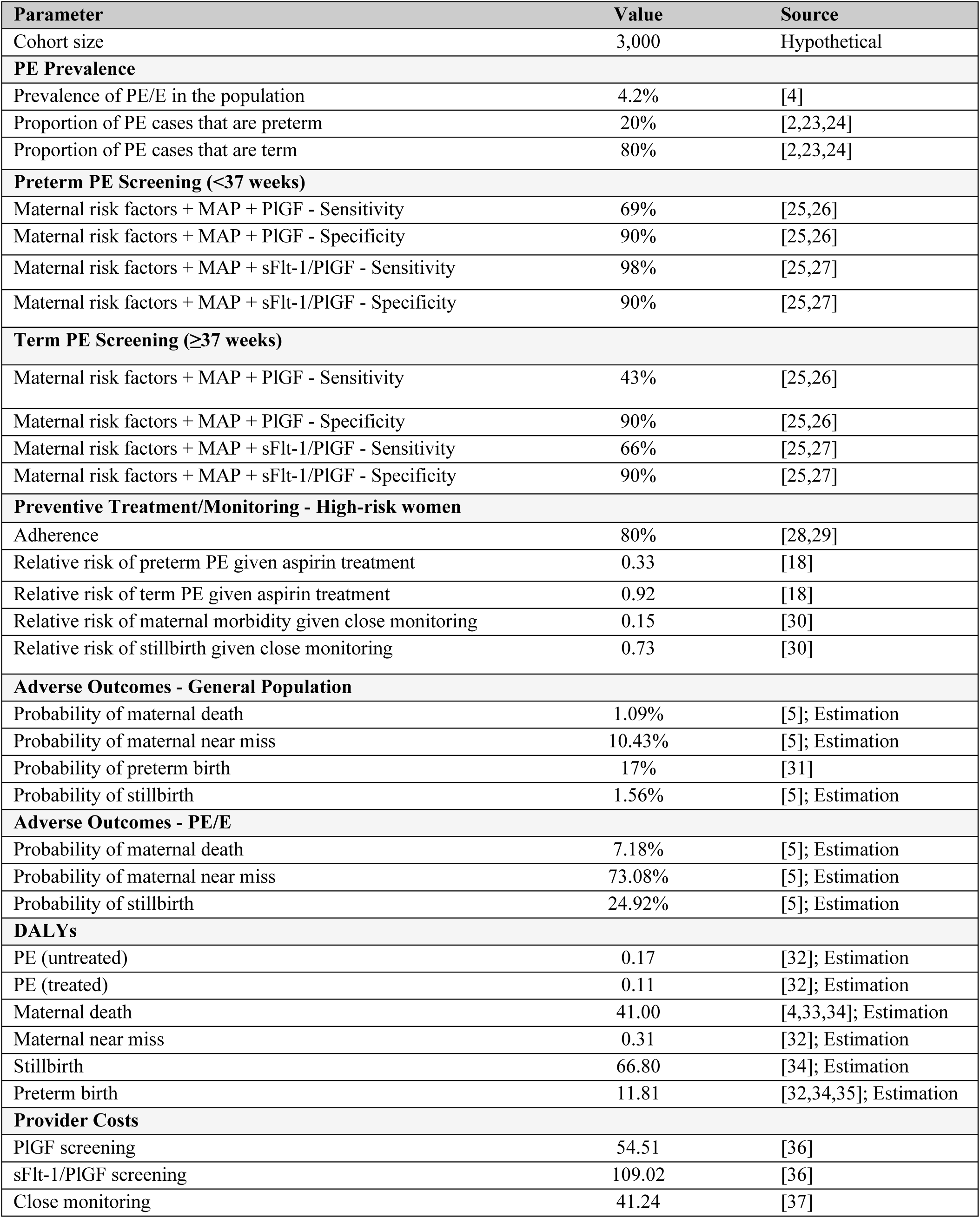
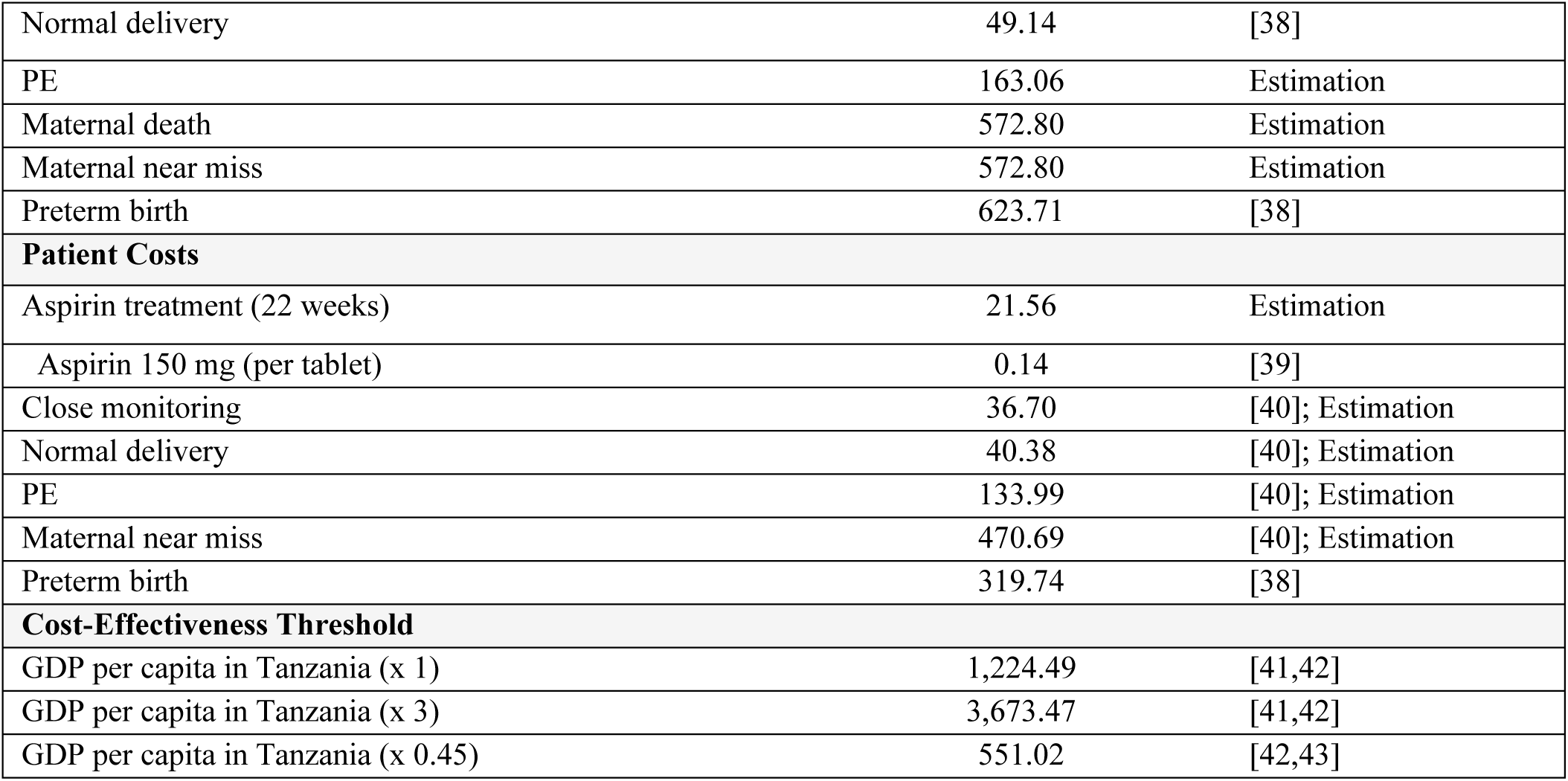
Data inputs. All costs are US$ at 2023 values. DALYs = Disability-adjusted life years. E = Eclampsia. GDP = Gross domestic product. MAP = Mean arterial pressure. PE = Pre-eclampsia. PlGF = Placental growth factor. sFlt-1 = Soluble fms-like tyrosine kinase-1.

| Parameter | Value | Source |
| --- | --- | --- |
| Cohort size | 3,000 | Hypothetical |
| **PE Prevalence** |  |  |
| Prevalence of PE/E in the population | 4.2% | [4] |
| Proportion of PE cases that are preterm | 20% | [2,23,24] |
| Proportion of PE cases that are term | 80% | [2,23,24] |
| **Preterm PE Screening (<37 weeks)** |  |  |
| Maternal risk factors + MAP + PlGF - Sensitivity | 69% | [25,26] |
| Maternal risk factors + MAP + PlGF - Specificity | 90% | [25,26] |
| Maternal risk factors + MAP + sFlt-1/PlGF - Sensitivity | 98% | [25,27] |
| Maternal risk factors + MAP + sFlt-1/PlGF - Specificity | 90% | [25,27] |
| **Term PE Screening (≥37 weeks)** |  |  |
| Maternal risk factors + MAP + PlGF - Sensitivity | 43% | [25,26] |
| Maternal risk factors + MAP + PlGF - Specificity | 90% | [25,26] |
| Maternal risk factors + MAP + sFlt-1/PlGF - Sensitivity | 66% | [25,27] |
| Maternal risk factors + MAP + sFlt-1/PlGF - Specificity | 90% | [25,27] |
| **Preventive Treatment/Monitoring - High-risk women** |  |  |
| Adherence | 80% | [28,29] |
| Relative risk of preterm PE given aspirin treatment | 0.33 | [18] |
| Relative risk of term PE given aspirin treatment | 0.92 | [18] |
| Relative risk of maternal morbidity given close monitoring | 0.15 | [30] |
| Relative risk of stillbirth given close monitoring | 0.73 | [30] |
| **Adverse Outcomes - General Population** |  |  |
| Probability of maternal death | 1.09% | [5]; Estimation |
| Probability of maternal near miss | 10.43% | [5]; Estimation |
| Probability of preterm birth | 17% | [31] |
| Probability of stillbirth | 1.56% | [5]; Estimation |
| **Adverse Outcomes - PE/E** |  |  |
| Probability of maternal death | 7.18% | [5]; Estimation |
| Probability of maternal near miss | 73.08% | [5]; Estimation |
| Probability of stillbirth | 24.92% | [5]; Estimation |
| **DALYs** |  |  |
| PE (untreated) | 0.17 | [32]; Estimation |
| PE (treated) | 0.11 | [32]; Estimation |
| Maternal death | 41.00 | [4,33,34]; Estimation |
| Maternal near miss | 0.31 | [32]; Estimation |
| Stillbirth | 66.80 | [34]; Estimation |
| Preterm birth | 11.81 | [32,34,35]; Estimation |
| **Provider Costs** |  |  |
| PlGF screening | 54.51 | [36] |
| sFlt-1/PlGF screening | 109.02 | [36] |
| Close monitoring | 41.24 | [37] |
| Normal delivery | 49.14 | [38] |
| PE | 163.06 | Estimation |
| Maternal death | 572.80 | Estimation |
| Maternal near miss | 572.80 | Estimation |
| Preterm birth | 623.71 | [38] |
| **Patient Costs** |  |  |
| Aspirin treatment (22 weeks) | 21.56 | Estimation |
| Aspirin 150 mg (per tablet) | 0.14 | [39] |
| Close monitoring | 36.70 | [40]; Estimation |
| Normal delivery | 40.38 | [40]; Estimation |
| PE | 133.99 | [40]; Estimation |
| Maternal near miss | 470.69 | [40]; Estimation |
| Preterm birth | 319.74 | [38] |
| **Cost-Effectiveness Threshold** |  |  |
| GDP per capita in Tanzania (x 1) | 1,224.49 | [41,42] |
| GDP per capita in Tanzania (x 3) | 3,673.47 | [41,42] |
| GDP per capita in Tanzania (x 0.45) | 551.02 | [42,43] |

Screening sensitivity and specificity are taken from research in HICs, assuming that locally derived PlGF and sFlt-1 thresholds will achieve comparable predictive performance. As the effectiveness of low-dose aspirin depends on compliance, we adopted the adherence reported in the ASPRE trial, the largest RCT on aspirin for PE prevention [28]. Similar adherence was observed in a Nigerian PE study, supporting its relevance in a comparable low-resource setting like Tanzania [29]. The same compliance rate is assumed for close monitoring, based on the expectation that patients respond similarly to preventive recommendations. In line with previous cost-effectiveness studies, the analysis considers only the effect of aspirin on PE onset. This approach isolates the role of preventive treatment in relation to the primary outcome of this analysis, as validated estimates of relative risk reduction for the broader range of adverse outcomes are unavailable. Commonly, preterm PE is associated with a higher rate of severe adverse outcomes than term PE [2,8]. However, consistent with its greater prevalence, von Dadelszen et al., (2022) found that term disease is associated with at least equivalent total numbers of maternal, and a significant proportion of perinatal adverse events compared with preterm PE. Based on these findings and the absence of outcome data disaggregated by gestational age, this model assumes equal risk of adverse outcomes for PE<37 and PE≥37. Probabilities of adverse maternal and neonatal outcomes, in both the presence and absence of PE were derived from the 2024 Quarterly Reports of the Bugando Medical Centre Department of Obstetrics and Gynaecology [5]. Since the reports did not provide the total number of PE/eclampsia cases but only those that resulted in MNMs, maternal deaths, and stillbirths, the total number of cases was approximated by multiplying the total number of deliveries by the prevalence of PE/eclampsia in the population (S2 Table).

Health outcomes were further quantified into disability adjusted life years (DALYs). As specific disability weights for PE, MNM, and preterm birth, are unavailable, we averaged weights from the Global Burden of Disease Study 2021 for related complications (S3 Table) [32]. Disability weights for PE vary by compliance with preventive treatment or close monitoring, with lower weights assigned to compliant women due to improved management and reduced likelihood of progression. DALYs for PE and MNM are computed by multiplying the disability weight by a one-year duration, capturing pregnancy, postpartum, and extended morbidity. DALYs for preterm birth account for severity of impairment, assuming lifelong duration equal to life expectancy at birth, and mortality (S4 Table). Maternal death DALYs equal years of life lost (YLL), calculated by subtracting mean age at PE diagnosis from female life expectancy in Tanzania. For stillbirth, YLL correspond to average life expectancy at birth for both sexes in Tanzania.

### Costs Estimation

All costs are presented in USD at 2023 values, adjusted using inflation calculators and currency converters (S5-S9 Table). Costs of PlGF and sFlt-1/PlGF screening include additional machine costs, reagents, service charges, training, and staff costs, and refer specifically to the Elecsys PLGF and sFlt-1 assays (Roche). Since MAP uses standard blood pressure readings, no additional costs are assumed. Aspirin cost per pregnancy was estimated by multiplying the unit cost by the duration of therapy. A daily dose of 150 mg was assumed, consistent with evidence that doses ≥100 mg reduce preterm PE risk [18]. Treatment duration was determined based on initiation at 15 weeks, aligning with evidence that aspirin is effective in reducing preterm PE risk when started before 16 weeks [18]. Close monitoring costs are shared between the provider, covering four additional ANC visits, and the patient, paying for urine tests at each visit and additional ultrasounds. ANC and postnatal care costs are omitted, as they are incurred equally across all screening options.

We assessed patient costs for three scenarios: a normal vaginal delivery, a vaginal delivery complicated by PE, and a MNM (S10 Table). For a normal vaginal delivery, we assumed discharge within 24 hours, consistent with regional norms. For the PE and MNM case, where costs can vary depending on the severity of the condition, we used data from randomly selected cases hospitalised for 7 and 13 days, respectively. PE-complicated and MNM deliveries were estimated to cost patients 3.32 and 11.66 times more than uncomplicated vaginal deliveries, respectively. Provider costs were derived by applying the same multipliers to baseline delivery costs, assuming similar scaling due to increased resource use. Provider costs of maternal death are assumed equal to MNM costs due to similar care intensity. Stillbirth and full-term neonates incur no additional costs beyond delivery. Preterm birth costs are proxied by provider costs for low-birth-weight neonates.

### Cost-effectiveness

The outcomes of this analysis are expressed in terms of incremental cost per DALY averted. The incremental cost-effectiveness ratio (ICER) is computed for each novel screening strategy with respect to current practice, and for Strategy 2 with respect to Strategy 1. We applied two approaches to determine intervention cost-effectiveness:

1. The WHO-CHOICE approach estimates a cost-effectiveness threshold (CET) based on a country’s gross domestic product (GDP) [41]. Interventions are “very cost-effective” if ICER < 1 x GDP per capita; “cost-effective” if ICER < 3 x GDP per capita. This analysis applies Tanzania’s 2023 GDP per capita ($1,224.49).
2. To counter noted limitations of the WHO-CHOICE approach, Woods et al., (2016) present country-level CETs based on the opportunity cost imposed on the country’s healthcare system to reflect its marginal productivity [43]. For Tanzania, the purchasing-power-parity-adjusted CET (2013 US$), was estimated as ranging between 4.7% - 95% of the country’s GDP per capita. An intervention is therefore “cost-effective” if ICER < 45% x GDP per capita.

### Sensitivity Analyses

To account for parameter uncertainty, we performed a probabilistic sensitivity analysis (PSA) using a Monte Carlo simulation with 2,000 iterations. We assigned Beta distributions to probability parameters and Lognormal distributions to relative risk parameters. For continuous, positive, right-skewed costs and DALYs data, a Gamma distribution was fitted. For DALYs associated with non-fatal conditions assumed to last up to one year, a Beta distribution was used. Where standard error estimates were not available, a coefficient of variation ranging from 0.2-0.3 was used to approximate uncertainty in the parameter values.

We conducted one-way sensitivity analyses to identify the parameters driving cost-effectiveness, varying each parameter within a range of ±30% from its baseline value.

### Scenario Analysis

We explored limiting biomarker-based screening to nulliparous women, given the expected financial challenges of a universal implementation in a low-resource setting. As maternal risk-factor-based screening relies on prior pregnancy history, it is less applicable to nulliparous women, making this sub-group a key target for new screening approaches. According to the latest TDHS-MIS, approximately 23% of pregnant women in Tanzania are nulliparous [15]. We applied this proportion in the model, assuming that only nulliparous women would be eligible for the new screening options – Strategy 1 and Strategy 2 – while the remaining would be subject to current practice. Unlike the base case analysis, we assumed that all high-risk women, regardless of parity, would receive preventive treatment and close monitoring, reflecting an improved standard of care with risk stratification and management following risk factor assessment. Additional data inputs utilised for performing this analysis are available in S2 Appendix.

## Results

### Base Case Analysis

The incremental cost per DALY averted associated with Strategy 1 and Strategy 2 as compared to current practice is $410.45 and $1,011.78, respectively (Table 2). Both Strategy 1 and 2 are deemed cost-effective under the WHO-CHOICE CET ($3,673.47), but only Strategy 1 remains cost-effective under the CET proposed by Woods et al., (2016) ($551.02). When compared to Strategy 1, Strategy 2 is dominated.

**Table 2.** Base-case analysis - Incremental cost per DALY averted. All costs are in US$ at 2023 values. DALYs = Disability-adjusted life years.

| Screening Strategy | Costs | DALYs | Incremental Cost per DALY averted |
| --- | --- | --- | --- |
| **Strategy 1/2 vs. current practice** |  |  |  |
| Current practice | 381.94 | 4.29 |  |
| Strategy 1 | 443.82 | 4.14 | 410.45 |
| Strategy 2 | 494.14 | 4.18 | 1,011.78 |
| **Strategy 2 vs. Strategy 1** |  |  |  |
| Strategy 1 | 443.82 | 4.14 |  |
| Strategy 2 | 494.4 | 4.18 | - 1,261.70 |

The possibility of early PE screening as provided for by Strategy 1, allows for an early initiation of aspirin, which is effective in reducing the risk of preterm PE [28]. In our analysis, approximately nine cases of preterm PE could be averted, resulting in an incremental cost per preterm PE case averted of $23,082 (Table 3). Only costs related to screening, preventive treatment, and monitoring were included in this analysis to isolate the impact of screening on preterm PE prevention rather than adverse outcomes.

**Table 3.** Base-case analysis - Incremental cost per PE<37 averted. Costs relate to screening, aspirin and close monitoring only, and are expressed in US$ at 2023 values. DALYs = Disability-adjusted life years. PE<37 = Preterm pre-eclampsia.

| Screening Strategy | Costs | DALYs | Incremental Cost per PE<37 averted |
| --- | --- | --- | --- |
| **Strategy 1 vs. current practice** |  |  |  |
| Current practice | - | 25.20 |  |
| Strategy 1 | 71.71 | 15.88 | 23,081.92 |

While neither strategy clearly improves maternal mortality, Strategy 2 is associated with the lowest number of MNMs, and Strategy 1 is the most effective in preventing stillbirths (Table 4).

**Table 4.** Adverse outcomes associated with each screening strategy. MNMs = Maternal near misses.

| Screening Strategy | Maternal Deaths | MNMs | Stillbirths |
| --- | --- | --- | --- |
| Current practice | 40.45 | 391.77 | 76.12 |
| Strategy 1 | 39.72 | 361.40 | 70.82 |
| Strategy 2 | 40.45 | 346.20 | 71.18 |

### Probabilistic Analysis

Fig 2 presents ICER distributions from 2,000 PSA iterations comparing biomarker-based strategies singularly with current practice and directly with each other. Strategy 1 consistently yielded higher costs and better outcomes compared to current practice, while Strategy 2 showed greater uncertainty, especially in terms of health outcomes, with a portion of the iterations dominated by current practice. The scatterplot confirmed that Strategy 2 is dominated by Strategy 1 in most iterations.

**Fig 2.**
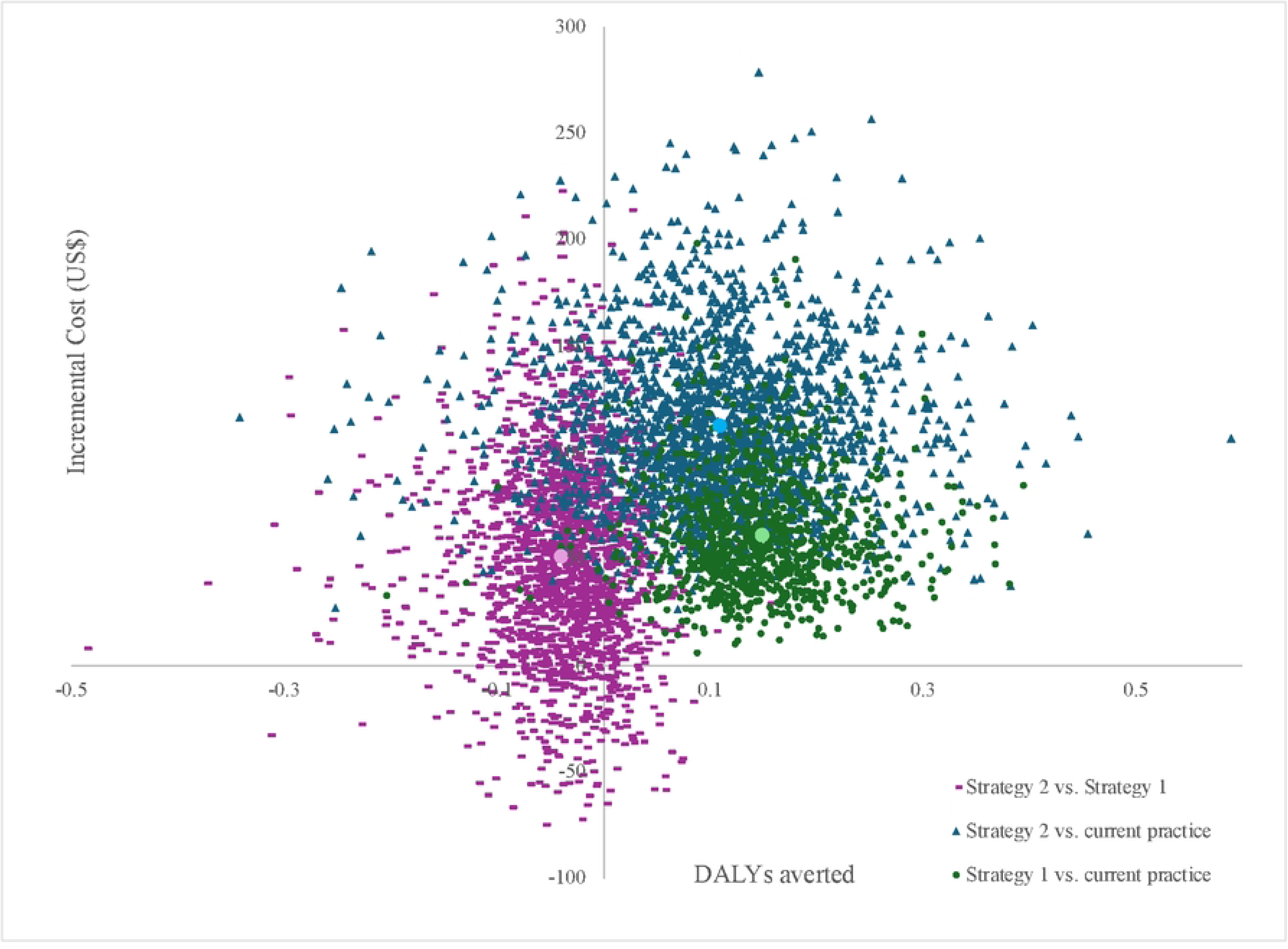
Distributions of ICERs for Strategy 1 vs. current practice, Strategy 2 vs. current practice, and Strategy 2 vs. Strategy 1. The bigger point estimates, marked in light purple, light green, and light blue represent the mean ICERs of each of the three distributions. Incremental costs are in US$ at 2023 values. DALYs = Disability-adjusted life years. ICERs = Incremental cost-effectiveness ratios.

The cost-effectiveness acceptability curves in Fig 3 show that once the CET exceeds approximately $410, Strategy 1 becomes more likely to be cost-effective compared to the other approaches, with a 65% and an 84% probability of being cost-effective at a CET of $551.02 and of $3,673.47, respectively. Strategy 2 is consistently dominated by Strategy 1 across all threshold values but becomes more favourable over current practice at CETs above $1,050.

**Fig 3.**
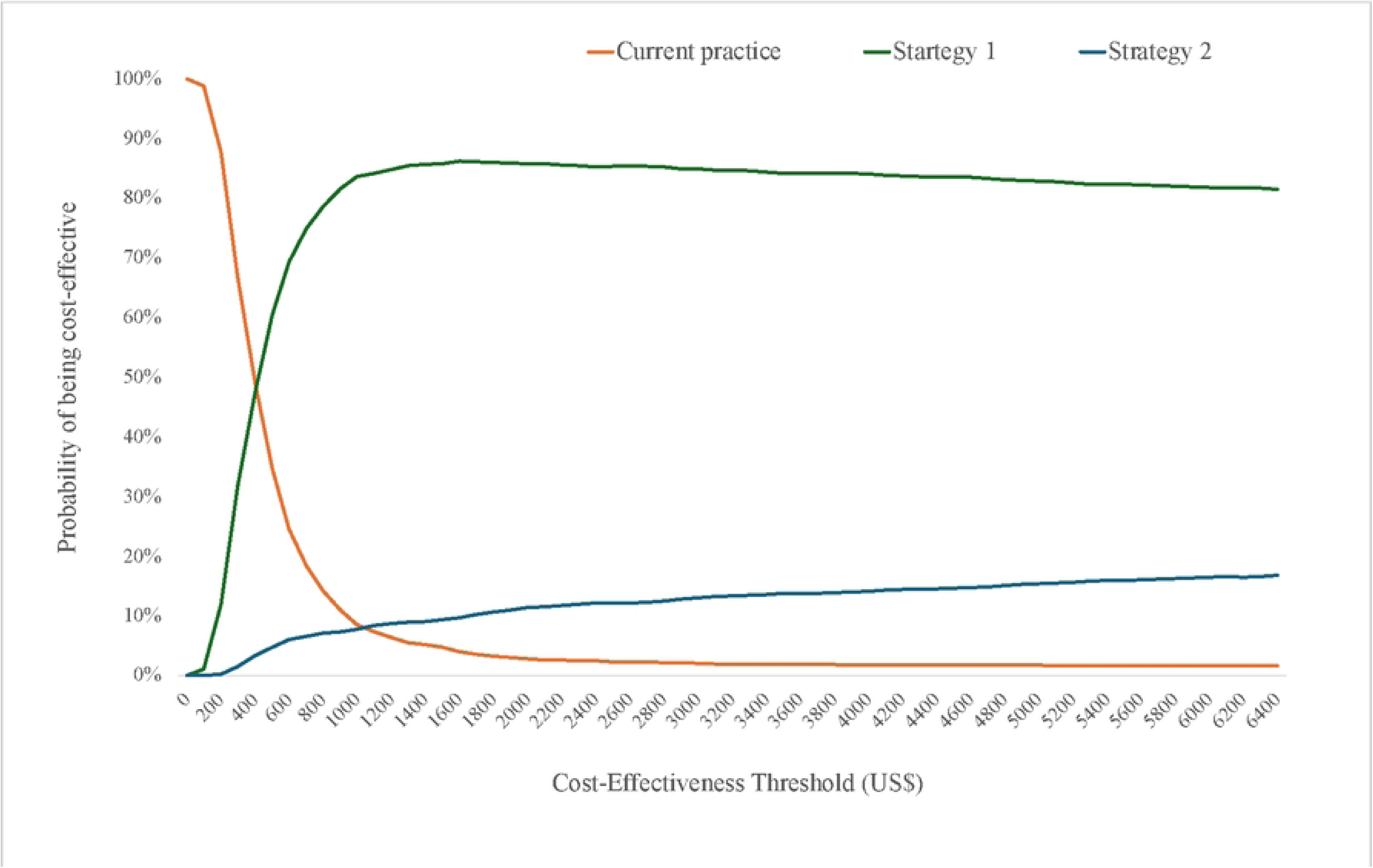
Cost-effectiveness acceptability curves for current practice, Strategy 1, and Strategy 2. Cost-effectiveness thresholds are in US$ 2023 values.

The cost-effectiveness acceptability frontier in Fig 4 shows that current practice is only viable at low CETs but is quickly outperformed by Strategy 1 as more resources become available. The absence of Strategy 2 suggests that the latter never yields the highest cost-effectiveness probability.

**Fig 4.**
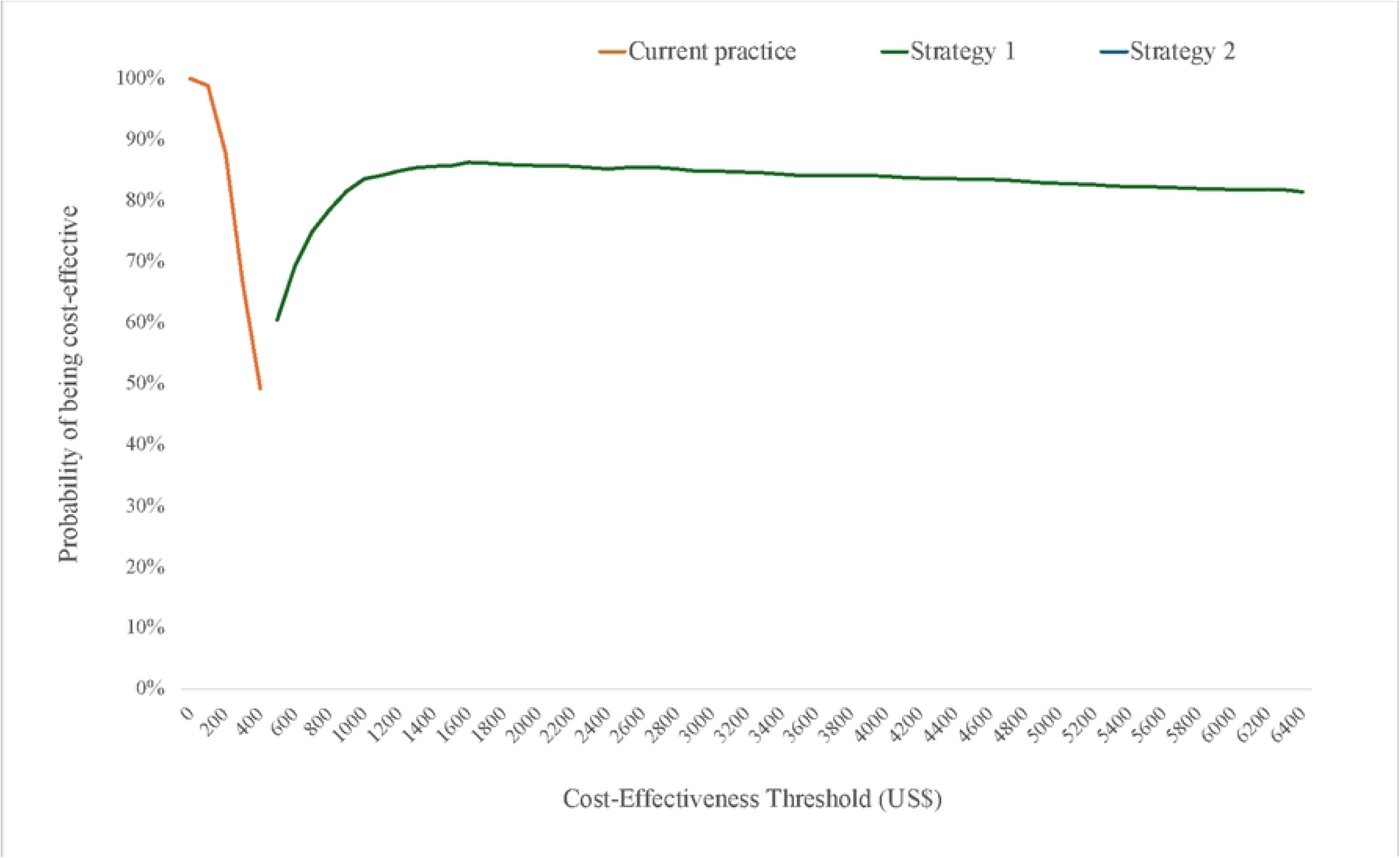
Cost-effectiveness acceptability frontier for current practice, Strategy 1, and Strategy 2. Cost-effectiveness thresholds are in US$ 2023 values.

### Sensitivity Analyses

When comparing Strategy 1 with current practice (Fig 5), the most influential parameter is the proportion of PE term cases. As Strategy 1 is particularly effective in preventing preterm PE through early aspirin initiation, the ICER increases with a greater incidence of term rather than preterm disease. Similarly, a lower prevalence of PE and reduced effectiveness of close monitoring elevate the ICER by diminishing the intervention’s overall health impact. Despite the acknowledged parameter uncertainty, the ICER remains below the WHO-CHOICE CET ($1,224.49), supporting the conclusion that Strategy 1 is very cost-effective. Under the CET proposed by Woods et al., (2016) ($551.02), Strategy 1 remains cost-effective except when the proportion of term cases is very high.

**Fig 5.**
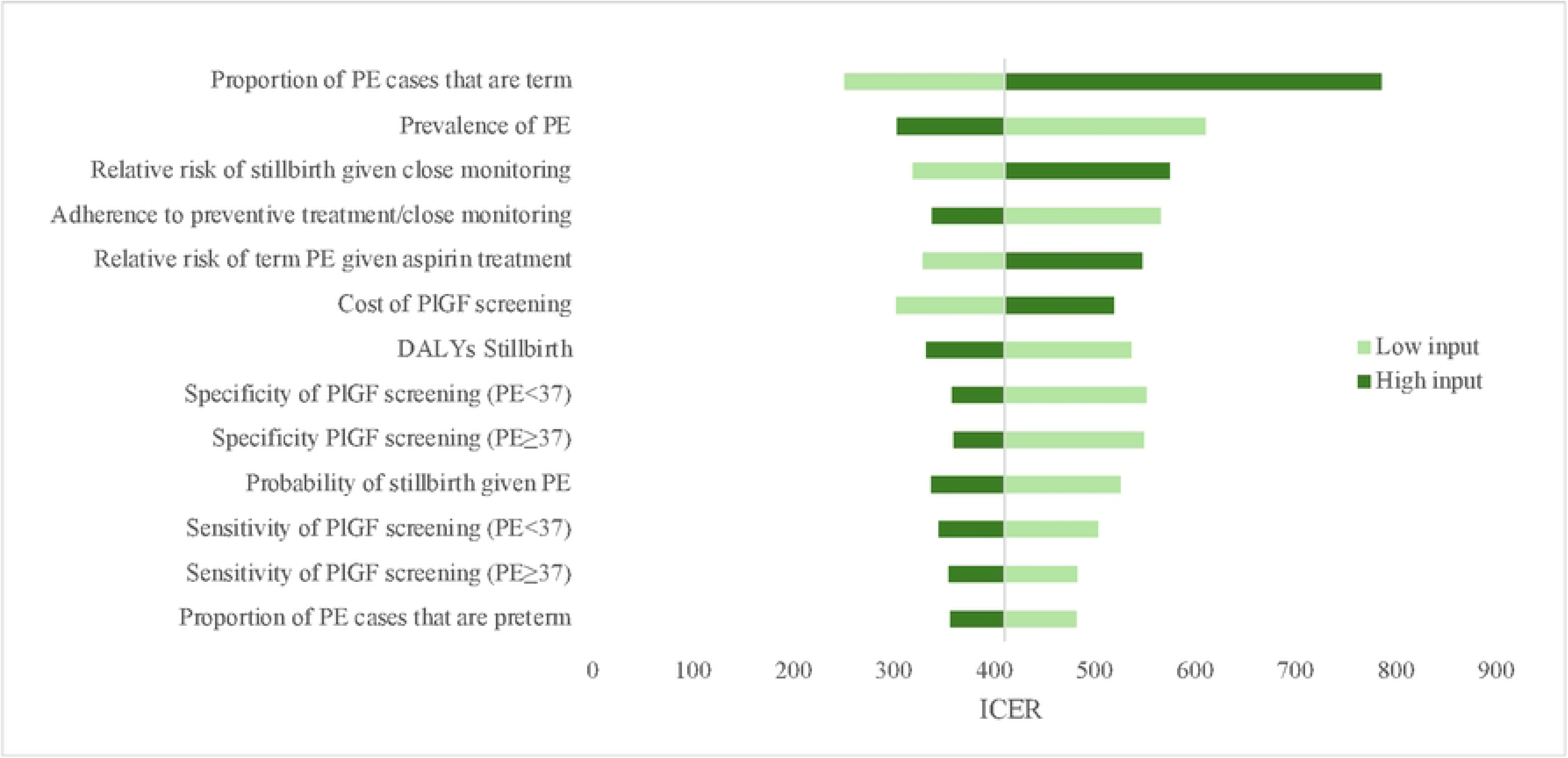
One-way sensitivity analyses. Tornado diagram for Strategy 1 vs. current practice. Input parameters are ranked from top to bottom according to the magnitude of their impact on the ICER. Each parameter is varied within a ±30% range of its baseline value. The ICER is expressed in terms of incremental 2023 US$ per DALY averted. DALYs = Disability-adjusted life years. ICER = Incremental cost-effectiveness ratio. PE = Pre-eclampsia. PE<37 = Preterm PE. PE≥37 = Term PE. PlGF = Placental growth factor.

The corresponding one-way sensitivity analysis for Strategy 2 vs. current practice is available in S1 Appendix, in addition to one-way sensitivity analyses of the most influential parameters going beyond the ±30% range.

### Scenario Analysis

When introducing biomarker-based PE screening only for nulliparous women, while strengthening current practice with risk stratification, preventive treatment and close monitoring for high-risk women, the incremental cost per DALY averted is $184.15 for Strategy 1 and $413.33 for Strategy 2. At the WHO-CHOICE CET ($1,224.49), both strategies are very cost-effective, and they remain cost-effective even at the CET proposed by Woods et al., (2016) ($551.02). Consistent with the main findings, Strategy 1 dominates Strategy 2 (S2 Appendix).

### Key Parameters for Data Collection

Table 5 lists key parameters along with suggested approaches for data collection during the planned PRESHA trial to enable a full and robust cost-effectiveness analysis of PE screening interventions. These parameters require particular consideration, as they have either shown considerable influence on the ICER, or relied on rough approximations due to data gaps.

**Table 5.**
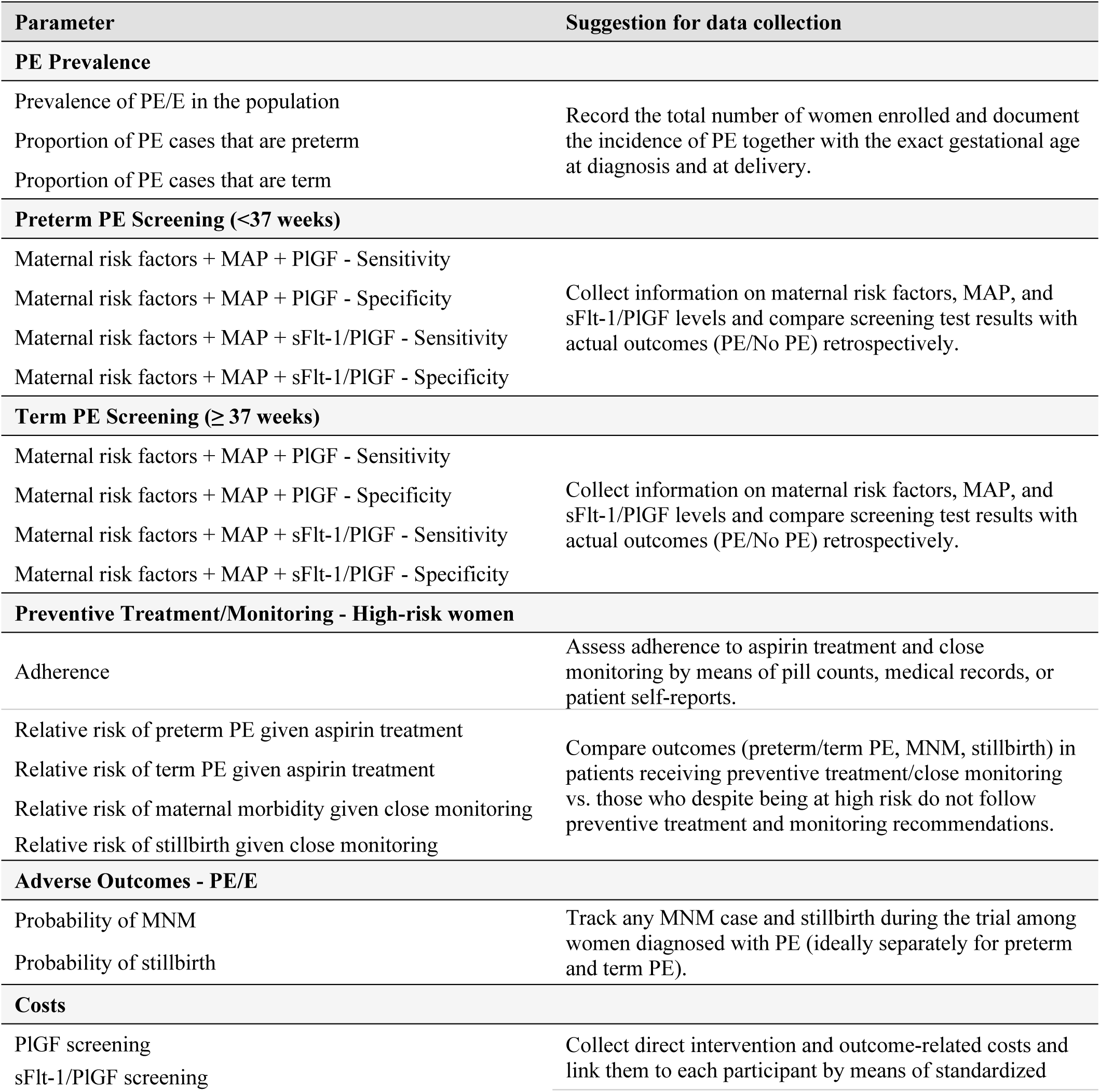
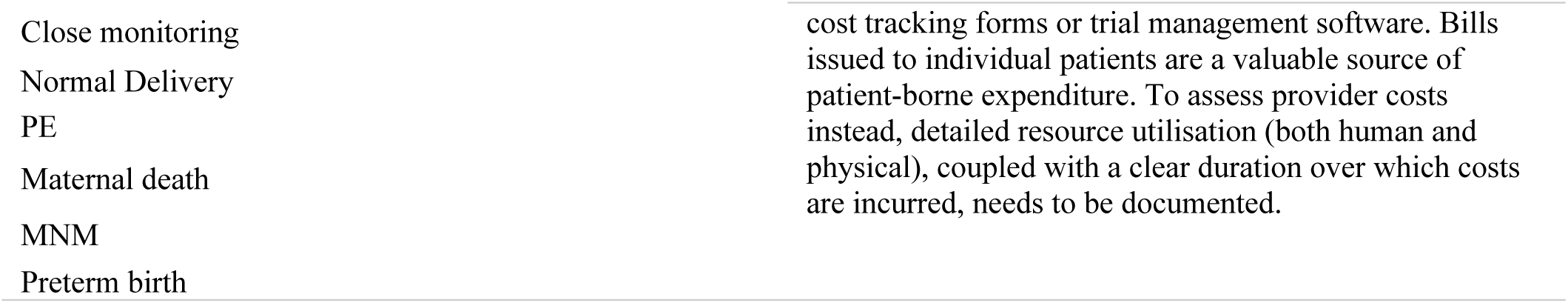
Key parameters and recommendations for trial data collection. DALYs = Disability-adjusted life years. E = Eclampsia. MAP = Mean arterial pressure. MNM = Maternal near miss. PE = Pre-eclampsia. PlGF = Placental growth factor. sFlt-1 = Soluble fms-like tyrosine kinase-1.

## Discussion

This study addresses a significant evidence gap by evaluating the cost-effectiveness of biomarker-based PE screening in a low-resource setting in sub-Saharan Africa. This pre-trial economic evaluation shows that supplementing current PE screening with biomarkers may be cost-effective in Tanzania. Particularly, Strategy 1, introducing PlGF, followed by aspirin prophylaxis and close monitoring for high-risk women, provided an ICER well below the applied CETs, aligning with evidence from HICs that early biomarker-based screening combined with aspirin prophylaxis and intensive monitoring effectively prevents preterm PE [11,19,20]. The cost per preterm PE case averted by Strategy 1 ($23,082), was consistent with estimates reported by Shmueli et al. (2012), Park et al. (2021), and Zakiyah et al. (2022), who evaluated comparable strategies in Israel, Australia, and European high-income countries, respectively. Other studies in HICs have also demonstrated that biomarker-based screening can be cost-saving, due to reduced hospitalisation and complication-related costs, further supporting our results [10,19,21].

Strategy 2, introducing the sFlt-1/PlGF ratio from mid-pregnancy, was cost-effective only under the WHO-CHOICE CET and dominated by Strategy 1. However, it may still represent a realistic opportunity of reducing adverse outcomes, as it is consistent with a later average initiation of ANC in low-income settings. To address local challenges, we explored a scenario in which nulliparous women undergo biomarker-based screening, while parous women are assessed by risk factors. This approach acknowledges financial and operational constraints and produces real-world evidence to inform the gradual expansion of the intervention to all pregnant women. At the same time, it highlights the need to strengthen existing practice to ensure optimal use of current tools and to improve outcomes irrespective of the adoption of advanced screening.

Our findings should be interpreted in light of several assumptions. Screening was modelled as occurring only at the first ANC visit, excluding repeat testing. Despite guidelines recommend combined aspirin and calcium supplementation for PE prevention, our analysis modelled prophylaxis with aspirin alone, potentially underestimating treatment effectiveness. Conversely, assuming high adherence may overstate effectiveness, given the cultural, educational, and financial barriers to adherence in LMICs [44].

The close monitoring framework was adapted from HICs and may not fully reflect the reality of Tanzania. Monitoring costs are likely underestimated, as hospitalisation and hypertensive medication were excluded due data limitations. Postpartum costs were omitted given their variability and limited relevance to this pre-trial evaluation.

Delivery costs were derived from individual billing records, as averaging across cases was unfeasible due to lack of data. Sensitivity analyses, however, showed limited impact of these estimates on cost-effectiveness outcomes. Finally, the short time horizon excludes long-term maternal and neonatal outcomes, likely underestimating the true benefits of screening.

Robust, evidence-based decisions on introducing biomarkers require more precise costs and outcomes estimates, achievable through improved data availability, reporting, and investment in locally relevant studies and trials. Longer time horizons would better capture long-term health and economic consequences of PE, enabling more realistic DALYs estimates and analyses from a broader societal perspective.

Finally, further research in LMICs is needed to adapt biomarker-based screening to low-resource contexts, focusing on point-of-care tests and community health workers engagement to enhance feasibility, as demonstrated in the CLIP trials [45]. Establishing locally appropriate biomarker thresholds is also critical, given population differences observed in previous studies [46].

While final policy recommendations should await the post-trial assessment of PRESHA, our analysis anticipates that early biomarker-based PE screening is likely cost-effective in Tanzania. Policymakers should consider phased integration into ANC, initially focusing on nulliparous women. Supportive educational policies for healthcare professionals and patients, may be developed in advance to facilitate successful implementation of a more comprehensive and cost-efficient PE screening.

## Data Availability

The data on which the research relies are all reported and/or appropriately referenced in the manuscript and supporting information document.

## Glossary

ANC: Antenatal care
CET: Cost-effectiveness threshold
DALYs: Disability-adjusted life years
E: Eclampsia
GDP: Gross domestic product
HICs: High-income countries
ICER: Incremental cost-effectiveness ratio
LMICs: Low- and middle-income countries
MAP: Mean arterial pressure
MNM: Maternal near miss
PE: Pre-eclampsia
PlGF: Placental growth factor
PRESHA: PREventing Severe Hypertensive Adverse events in pregnancy and childbirth
sFlt-1: Soluble fms-like tyrosine kinase-1
TDHS-MIS: Tanzania Demographic and Health Survey and Malaria Indicator Survey
YLL: Years of life lost

## Supporting information

### Tables

**S1 Table. CHEERS 2022 Checklist.** Source: Husereau et al., 2022.

**S2 Table. Probability of adverse pregnancy outcomes for women without/with PE/E**. E = Eclampsia. MNM = Maternal near miss. PE = Pre-eclampsia.

**S3 Table. Disability weights associated with PE, MNM, and preterm birth.** ª Moderate impairments with a disability weight of 0.24, which is closer to the weights assigned to severe impairments, were grouped with the severe category, while moderate impairments with disability weights closer to mild impairment were categorised as mild impairments.

**S4 Table**. **DALYs associated with preterm birth**. DALYs = Disability-adjusted life years.

**S5 Table. Cost of aspirin treatment for women at high risk of PE, from 15 to 36 weeks of gestation.** Source: Bugando Medical Centre Pharmacy, 2025. Exchange rate as of March 6th, 2025: 1 TZS = 0.000382719 USD; 1 US$ = 2,612.88 TZS. Cumulative inflation rate (2025 US$ – 2023 US$): -4.1%. TZS = Tanzanian Shilling.

**S6 Table. Cost of the tests for the measurement of biomarker levels**. Source: NICE, 2022. Exchange rate as of March 6th, 2025: 1 GBP = 1.28934 USD; 1 US$ = 0.775591 GBP. Cumulative inflation rate (2025 US$ – 2023 US$): -4.1%. GBP = Great British Pound. PlGF = Placental growth factor. sFlt-1 = Soluble fms-like tyrosine kinase-1.

**S7 Table. Cost of close monitoring for women at high risk of PE.** Sources: Definition of close monitoring by Zakiyah et al., 2022; ANC visit cost retrieved from Chamani et al., 2021; cost of ultrasound and urine analysis from billing records from Bugando Medical Centre, 2025. Exchange rate as of March 6th, 2025: 1 TZS = 0.000382719 US$; 1 US$= 2,612.88 TZS. Cumulative inflation rate (2018 US$ – 2023 US$): 21.3%; Cumulative inflation rate (US$ 2025 – US$ 2023): -4.1%. ANC = Antenatal care. TZS = Tanzanian Shilling.

**S8 Table. Provider costs for an uncomplicated vaginal delivery, a delivery complicated by PE, a delivery complicated by MNM/maternal death, and preterm birth.** Source: Mori et al., 2020. Costs associated with PE and MNM/MD were approximated by multiplying the cost of a vaginal delivery ($49.14) by the respective multiplier computed in the main analysis: 3.32 for PE and 11.66 for MNM/MD. Preterm birth costs were approximated by the costs of manging LBW neonates ($623.71) based on comparable resource use. Cumulative inflation rate (2018 US$ – 2023 US$): 21.3%. LBW = Low birth weight. MD = Maternal death. MNM = Maternal near miss. PE = Pre-eclampsia.

**S9 Table. Patient costs for a preterm birth.** Source: Mori et al., 2020. Preterm birth costs were approximated by the costs of delivering LBW neonates ($319.74) based on comparable resource use. Cumulative inflation rate (2018 US$ – 2023 US$): 21.3%. LBW = Low birth weight.

**S10 Table. Direct patient costs associated with a normal, PE-complicated, and MNM delivery.** Cost ratios indicate the incremental cost of a PE-complicated and MNM-complicated delivery with respect to an uncomplicated delivery and are expressed in terms of multipliers. The following criteria are used for conversion into US$ 2023 values: Exchange rate as of March 6th, 2025: 1 TZS = 0.000382719 US$; 1 US$ = 2,612.88 TZS; Cumulative rate of inflation (2025 US$ - 2023 US$): -4.1%. MNM = Maternal near miss. TZS = Tanzanian Shilling.

### Appendices

#### S1 Appendix - Sensitivity Analysis

**S1 Fig. One-way and two-way sensitivity analyses of the most influential parameters on the ICER of Strategy 1 vs. current practice.** For assessing the impact on the ICER of the variation in the proportion of PE cases that are term, a two-way sensitivity analysis is needed, as this proportion changes simultaneously with the complementary proportion of PE cases that are preterm. The ICER is expressed in terms of incremental cost (US$ 2023) per DALY averted. ICER = Incremental cost-effectiveness ratio. pAdh = Adherence to preventive treatment/close monitoring. pPrev = Prevalence of PE. pPrev_PPE = Proportion of PE cases that are preterm. pPrev_TPE = Proportion of PE cases that are term. RR_Stillbirth_Monitoring = Relative risk of stillbirth given close monitoring.

**S2 Fig. One-way sensitivity analysis. Tornado diagram for Strategy 2 vs. current practice.** Input parameters are ranked from top to bottom according to the magnitude of their impact on the ICER. Each parameter is varied within a ±30% range of its baseline value. The ICER is expressed in terms of incremental 2023 US$ per DALY averted. DALYs = Disability-adjusted life years. ICER = Incremental cost-effectiveness ratio. MNM = Maternal near miss. PE = Pre-eclampsia. PE<37 = Preterm PE. PE≥37 = Term PE. PlGF = Placental growth factor. sFlt-1 = Soluble fms-like tyrosine kinase-1.

**S3 Fig. One-way and two-way sensitivity analyses of the most influential parameters on the ICER of Strategy 2 vs. current practice.** The ICER is expressed in terms of incremental cost (US$ 2023) per DALY averted. DALY = Disability-adjusted life years. DALY_Stillbirth = DALYs associated with stillbirth. ICER = Incremental cost-effectiveness ratio. pPrev = Prevalence of PE. pStillbirth_PE = Probability of stillbirth given PE. RR_Stillbirth_Monitoring = Relative risk of stillbirth given close monitoring.

#### S2 Appendix - Scenario analysis

**S11 Table. Additional data inputs required to perform the scenario analysis.** Strategy 1 occurring before 16 weeks of gestation is associated with maternal risk factor screening in early pregnancy (11-13 weeks), while Strategy 2 taking place after 24 weeks is associated with late pregnancy maternal risk factor screening (30-34). PE = Pre-eclampsia.

**S12 Table. Scenario analysis – Incremental cost per DALY averted.** All costs are in US$ at 2023 values. DALYs = Disability-adjusted life years.

S13 Table. Scenario analysis – Difference in adverse outcomes and preterm PE cases averted with respect to the base case analysis. All costs are in US$ at 2023 values. MNMs = Maternal near misses. PE<37 = Preterm pre-eclampsia.

